# Smartphone-connected, battery-free aptamer-based digital test for quantification of procalcitonin and monitoring of infection at the point of care

**DOI:** 10.64898/2026.07.27.26358798

**Authors:** Yuanjun Cai, Jose M. R. Flauzino, Abdulkadir Sanli, Tinghao Hu, Hong Seok Lee, Alex Silva-Collins, Laura Gonzalez-Macia, Suzanne Williams, Stefania Frederico, Richard C. Wilson, Timothy M. Rawson, Firat Güder

## Abstract

**Background:** Rapid and accurate measurement of procalcitonin (PCT) is useful for diagnosing bacterial infections and guiding antibiotic therapy, yet current laboratory-based immunoassays require centralised infrastructure, delaying clinical decision-making and limiting access in low-resource settings. We developed a battery-free, smartphone-connected electrochemical lateral flow assay with linked analytics for the detection of PCT (ELLA-PCT) enabling quantitative testing at the point of care without conventional laboratory instrumentation.

**Methods:** We designed a competitive electrochemical lateral flow assay using gold nanoparticles co-functionalised with a PCT-specific DNA aptamer and ferrocene hexanethiol as a redox reporter. A miniaturised near-field communication (NFC) potentiostat embedded within a disposable cassette enabled wireless electroanalytical measurements using a smartphone. Analytical performance was assessed in buffer and serum, including limit of detection (LOD), linearity, specificity, and stability. Clinical evaluation was performed on 27 serum samples from nine adults undergoing antibiotic treatment for suspected bacterial infection, with results compared against the reference Time-Resolved Amplified Cryptate Emission (TRACE) assay.

**Findings:** ELLA-PCT achieved an LOD of 46 pg/mL and a linear detection range of 0.5–100 ng/mL. The ELLA-PCT assays remained stable for three months under ambient storage conditions, and cross-reactivity with calcitonin, C-reactive protein, and interleukin-6 remained below clinically relevant thresholds. Clinical results showed strong correlation with TRACE (R² = 0·979, p < 0·0001), with a mean bias of 0·05 ng/mL and narrow 95% limits of agreement (– 0·69 to 0·89 ng/mL). Importantly, ELLA-PCT delivered results in only 30 minutes without requiring external power or clinical laboratory instrumentation. In comparison to TRACE, the total turnaround time was reduced by at least 50%, which typically requires at least one hour from sample collection to results at a hospital setting.

**Interpretation:** ELLA-PCT is an antibody-free, smartphone-connected test that provides laboratory-grade quantitative PCT measurement using only an NFC-enabled electroanalytical sensor and a mobile phone. The platform has a strong potential to decentralise diagnosis of infectious diseases, support antibiotic stewardship, and enable remote, real-time monitoring in outpatient and resource-limited settings. Larger studies are needed to evaluate the use of whole-blood samples and the integration of the platform into digital clinical workflows.

**Funding:** The authors acknowledge the Gates Foundation (Grand Challenges Explorations scheme under grant numbers OPP1212574 and INV-038695) for financial support. FG and JMRF acknowledge Research England International Science Partnerships Fund (References: G08259 and G08288) and the Biotechnology and Biological Sciences Research Council Impact Acceleration Account (Reference: PSR417). FG and AS acknowledge Innovate UK BMC (Reference: PA2131). FG and LGM acknowledge funding from the European Union’s Horizon 2020 research and innovation programme under the Marie Sklodowska-Curie grant agreement No 101025390. FG acknowledge the Bezos Earth Fund through the Bezos Centre for Sustainable Protein (BCSP/IC/001). TMR, RW, SF, & SW acknowledge The Wellcome Trust funded Centres for Antimicrobial Optimisation Network (CAMO-NET. Reference: 226691/Z/22/Z)

## Introduction

Sepsis remains a leading cause of mortality worldwide, and early diagnosis can be critical to patient survival.^1^ Procalcitonin (PCT), a peptide precursor of calcitonin, is normally produced at low concentrations by thyroid C cells, but during systemic bacterial infection its circulating concentration increases substantially, largely through extra-thyroidal production in multiple tissues.^2^ Compared with conventional inflammatory markers such as C-reactive protein (CRP), PCT concentrations increase more rapidly in response to systemic bacterial infection, becoming detectable within 2–6 h and generally peaking within 12–24 h.^3^ Following effective treatment and control of the infection, PCT concentrations decline with a half-life of approximately 24 h, whereas persistently elevated or increasing concentrations may indicate ongoing infection or an inadequate response to treatment.^4^ These characteristics make PCT a valuable biomarker for diagnosing bacterial infections, assessing disease severity, and guiding antibiotic therapy.^5^ Recent randomised clinical trials further support the clinical relevance of PCT-informed care pathways: rapid PCT testing combined with NEWS2 assessment was associated with reduced 28-day mortality in patients managed for suspected sepsis in emergency departments, while daily PCT-guided monitoring in critically ill patients with suspected sepsis safely reduced antibiotic duration compared with standard care.^6,7^

Repeated measurements of PCT during the evaluation and treatment of suspected bacterial infections can be limited by the need for centralised laboratory infrastructure, specialised equipment, and trained personnel. These constraints may delay the availability of results needed to support timely treatment decisions. There is, therefore, a need for portable, reliable, and cost-effective systems that enable healthcare professionals to measure PCT closer to the point of care and facilitate the monitoring and differentiation of bacterial infections. The gold-standard method for measuring PCT in blood is Time-Resolved Amplified Cryptate Emission (TRACE).^8^ Despite being highly sensitive, TRACE requires complex instrumentation, lengthy processing times, and the need for highly trained laboratory personnel. Other techniques, such as enzyme-linked fluorescent assays^9,10^ electrochemiluminescence immunoassays,^11,12^ and others ^13–16^ also suffer from similar shortcomings as TRACE, limiting their use to centralised and well-resourced laboratories that do not demand rapid processing times.

Lateral flow assays (LFAs) are a powerful tool for point-of-care diagnostics due to their simplicity, rapid turnaround time, and low cost. Despite the clear advantages of LFAs as a diagnostic method, LFAs produce qualitative or at best semi-quantitative results, relying on visual interpretation of colourimetric signals which limit their performance and utility.^17^ LFAs have, therefore, historically been seen as a quick method to produce a “yes/no” type result which is only useful in directly identifying a pathogen.^18^ If the analytical target requires higher sensitivity or quantification, LFAs, in their current form, are unsuitable as a diagnostic approach.^19^

To address the limitations of conventional LFAs, we previously developed electrochemical lateral flow assay technologies integrating low-power readout and near-field communication (NFC) connectivity.^20,21^ More recently, we reported an electrochemical lateral flow assay with linked analytics (ELLA) for the surveillance of cassava brown streak disease in East Africa. ^22^ Although this platform was initially developed for agricultural diagnostics, its ability to provide quantitative measurements in a low-cost, portable, and battery-free format makes it suitable for translation to clinically relevant biomarkers. ELLA also employed antibodies as the biorecognition elements, but the cost of manufacturing and storage requirements of antibodies can restrict their reach as a diagnostic tool. In the present study, we used DNA aptamers, single-stranded DNA molecules selected for their high affinity and stability, as the biorecognition elements for the detection of PCT.^23^ The aptamers were conjugated to gold nanoparticles (AuNPs), and ferrocene hexanethiol was incorporated as a redox-active reporter to generate a quantitative electrochemical signal without fundamentally changing the established format, manufacture, or operation of an LFA. We evaluated an ELLA- PCT platform in controlled experiments using buffer and serum and subsequently tested it with serum samples from patients undergoing antibiotic treatment for suspected bacterial infection. ELLA-PCT measurements were compared with those obtained using TRACE as the clinical reference method.

## Methods

### Study design and reporting guidelines

We evaluated ELLA-PCT as a diagnostic accuracy and method-comparison study comprising analytical validation in buffer and serum, followed by clinical evaluation using serum samples from adult patients receiving intravenous antibiotics for suspected bacterial infection. ELLA- PCT was assessed as the index test and compared with time-resolved amplified cryptate emission (TRACE), the hospital reference method for PCT measurement. The study is reported in accordance with STARD guidelines for diagnostic accuracy studies. The ELLA-PCT assay working principle is illustrated in Fig. 1. The detailed methods are written in the supplementary information file.

**Figure 1.**
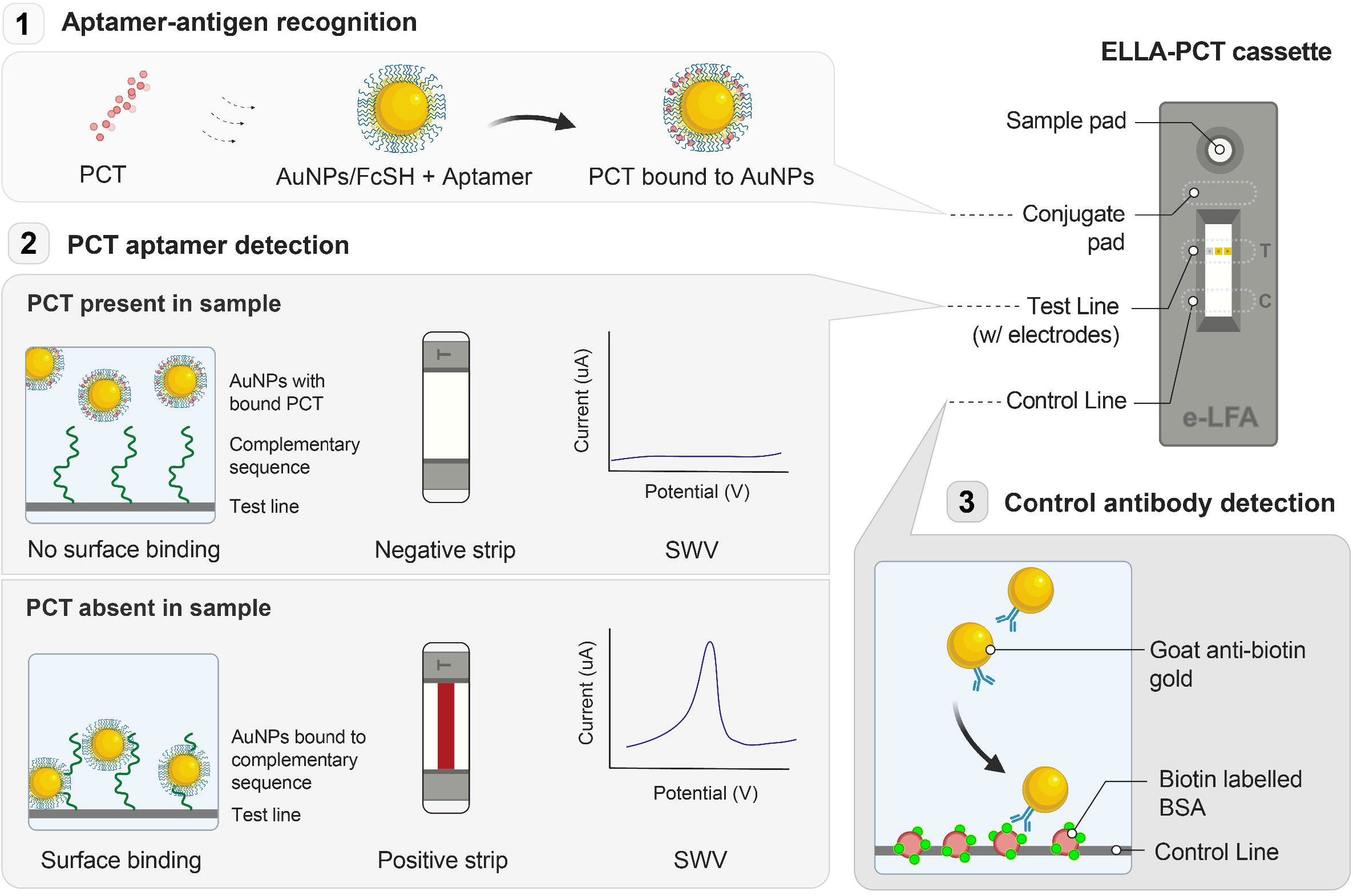
Working principle of the ELLA-PCT. The ELLA-PCT operates as a competitive assay in which sample flows across the lateral flow strip, enabling binding interactions between PCT and aptamer–AuNP conjugates (1). In the absence of PCT, AuNP–aptamer–ferrocene conjugates hybridise with complementary oligonucleotides on the test line, generating a colourimetric and electrochemical signal. Increasing PCT concentrations prevent this hybridisation, resulting in a proportional reduction in signal (2). For the control line, an anti-biotin nanoparticle binds to the biotinylated BSA printed in the control line (3).

### Materials and reagents

A PCT-specific DNA aptamer (51 nt) reported previously, was synthesised with a 5′-terminal thiol to enable attachment to AuNPs.^24^ Ferrocene hexanethiol (FcSH) was used as the redox reporter. Nitrocellulose strips, conjugate pads, sample pads, and absorbent pads were assembled into lateral flow cassettes designed to interface with a miniaturised NFC potentiostat (**Fig. S1**).

### Fabrication of aptamer-AuNP redox conjugates

AuNPs (40 nm) were functionalised through sequential conjugation of the thiolated aptamer and FcSH (**Fig. 2)**. Optimal aptamer loading was determined using a salt-induced aggregation assay^25^, identifying MOPS buffer (pH 7·0) and 48 µg/mL of aptamer as the optimal conditions (**Fig. S2**). Successful conjugation of FcSH and aptamer was confirmed using UV-visible spectroscopy, which demonstrated characteristic red-shifts and peak broadening consistent with increased surface functionalisation^26^ (**Fig. S3A**). Electrochemical activity of the conjugates was assessed by drop-casting functionalised nanoparticles onto screen-printed gold electrodes and performing square-wave voltammetry (SWV)^27^, with ferrocene carboxylic acid (FcCOOH) included as a non-thiolated control (**Fig. S3B**).

**Figure 2.**
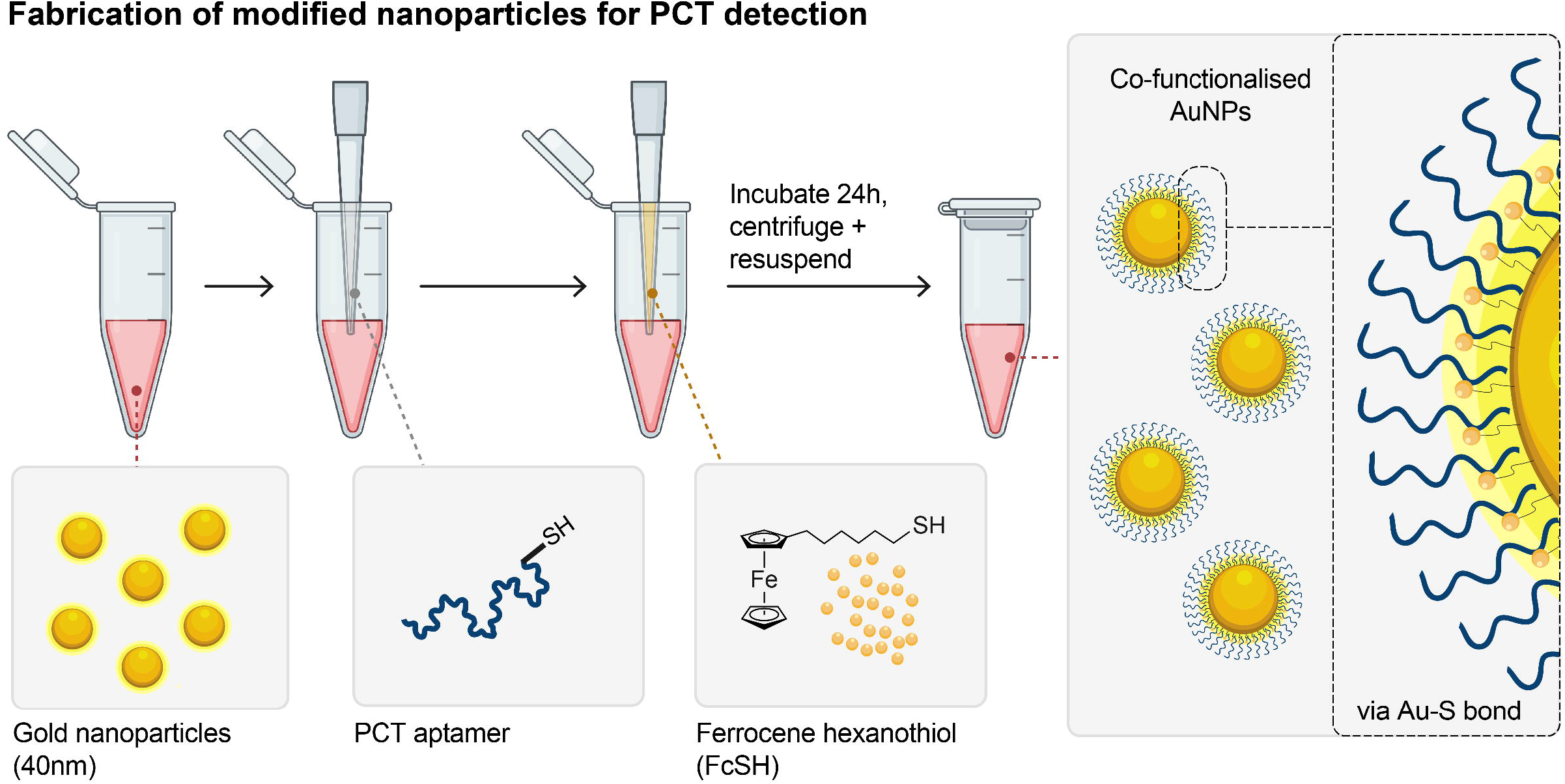
Fabrication of the signal-generating nanoparticles used in ELLA-PCT. Gold nanoparticles were modified with two components: a PCT-specific DNA aptamer, which acts as the recognition element for procalcitonin, and ferrocene hexanethiol (FcSH), which acts as an electrochemical reporter. The aptamer enables selective interaction with PCT, whereas ferrocene generates a measurable current when the strip is read by the NFC-powered sensor. Together, these modified nanoparticles convert PCT binding into a quantitative electrochemical signal, allowing PCT concentrations to be measured without conventional laboratory instrumentation.

### ELLA-PCT device architecture

Test strips were housed in a 3D-printed cassette incorporating a disposable printed circuit board (PCB) containing a three-electrode acupuncture-needle array (gold-plated working and counter electrodes; silver-plated reference electrode) (**Fig. 3A**). The PCB integrated an NFC potentiostat (SIC4341 from Silicon Craft Technology PLC, Thailand) enabling wireless signal acquisition and data transfer to an Android smartphone application (**Fig. 3B**). Electrodes were inserted post-assay development to avoid interference with sample flow dynamics.

**Figure 3.**
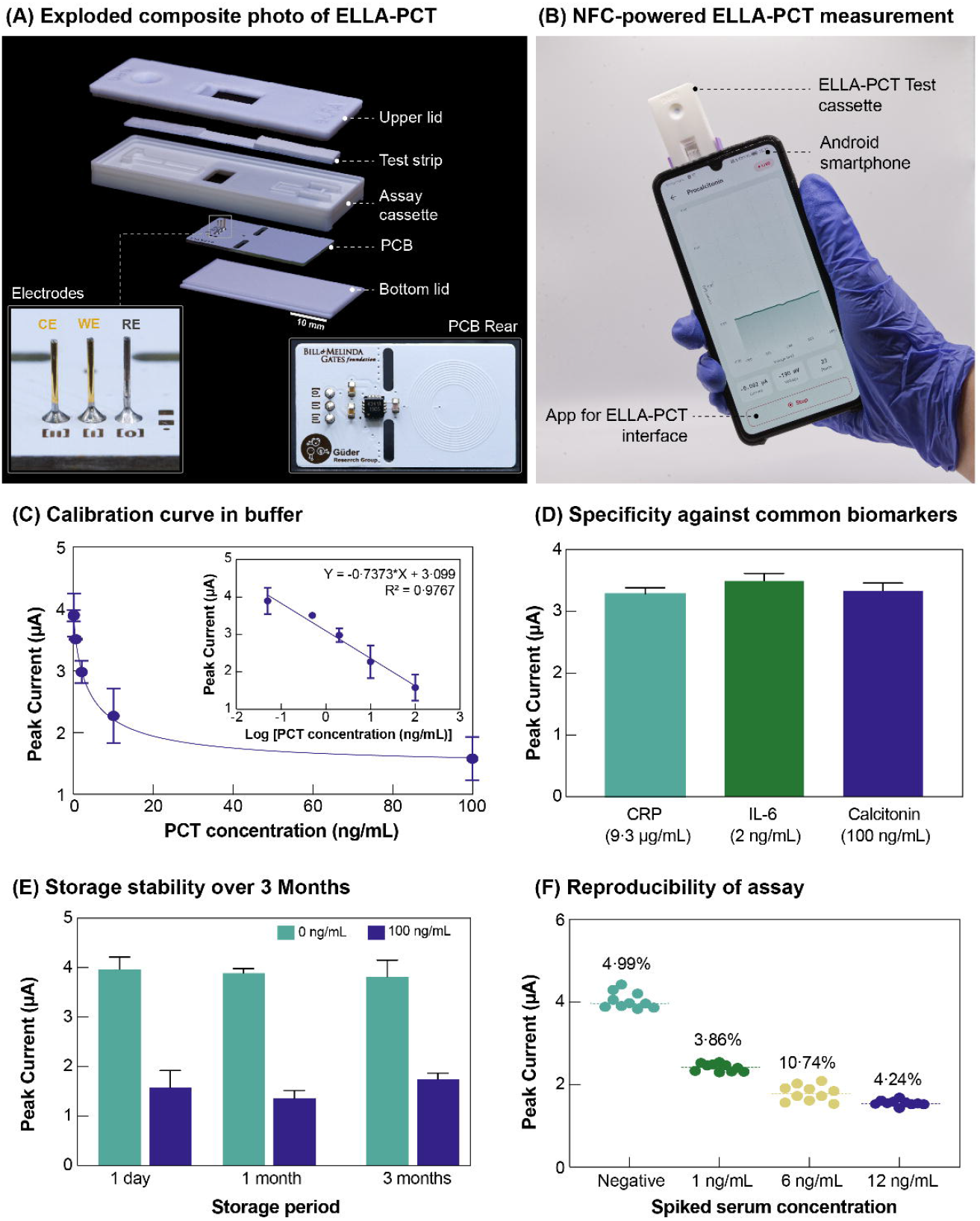
ELLA-PCT device architecture and analytical performance. **(A)** Exploded composite photograph of the ELLA-PCT disposable cassette, showing the upper lid, lateral flow test strip, assay cassette, printed circuit board (PCB), and bottom lid. The inset shows the three-electrode acupuncture-needle array used for on-strip electrochemical measurement, comprising counter, working, and reference electrodes. **(B)** Photograph of the NFC-powered ELLA-PCT system during measurement. The disposable cassette is wirelessly powered and read by an Android smartphone, which displays the electrochemical response through the ELLA-PCT application. **(C)** Calibration curve obtained in buffer, showing the inverse relationship between PCT concentration and peak current. Because ELLA-PCT uses a competitive assay format, higher PCT concentrations reduce nanoparticle capture at the test line and therefore decrease the measured electrochemical signal. The inset shows the linear relationship between peak current and the logarithm of PCT concentration (n = 3). **(D)** Specificity assessment against common inflammatory and endocrine biomarkers, including CRP, interleukin-6 (IL-6), and calcitonin. **(E)** Storage stability of the assay after 1 day, 1 month, and 3 months under ambient conditions, evaluated using negative samples and samples containing 100 ng/mL PCT. **(F)** Reproducibility of ELLA-PCT measurements in spiked serum samples across clinically relevant PCT concentrations, with coefficients of variation shown above each group.

### Electrochemical measurement protocol

Following sample addition, strips were incubated at ambient temperature. Needle electrodes were inserted after 30 minutes of sample running, which was determined to be the optimal measurement time point following evaluation of signal stability between 10 and 35 minutes (**Fig. S4**). SWV was performed directly on the test line to quantify the ferrocene (Fc/Fc ) redox current proportional to PCT-unbound AuNP–aptamer–FcSH conjugates.

### Analytical validation

Serial dilutions of recombinant human PCT (0–100 ng/mL) were prepared in buffer and serum. The limit of detection (LOD) was calculated as mean blank signal minus three standard deviations. Specificity was assessed using supraphysiological concentrations of potential interferents (calcitonin, CRP, IL-6). Long-term stability was evaluated under ambient storage for up to 3 months.

### Clinical samples and participants

We analysed 27 serum samples collected from nine adult patients receiving intravenous antibiotics for suspected bacterial infection (basic demographic and clinical characteristics were in the SI). Participants were recruited during pharmacokinetic steady state, and three samples were collected per participant at predefined timepoints relative to the antibiotic dosing interval: peak, midpoint, and trough. Patients received either meropenem or piperacillin/tazobactam according to their clinical treatment plan. The samples spanned clinically actionable PCT ranges for infection assessment and antibiotic-stewardship decision-making. Ethics approval was obtained from the London–Brighton and Sussex Research Ethics Committee (REC ref 22/LO/0063). Written informed consent was obtained from all participants.

### Index test and reference standard

ELLA-PCT was evaluated as the index test. The index test result was defined as the PCT concentration calculated from the electrochemical peak current using the serum calibration curve. TRACE was used as the reference standard for comparison. ELLA-PCT and TRACE measurements were compared using the same serum samples. ELLA-PCT measurements were performed before comparison with TRACE results, and assay operators were not involved in routine clinical decision-making.

### Statistical analysis

Analytical standard curves were fitted using a four-parameter logistic model. Agreement between the ELLA-PCT and TRACE was evaluated using Bland–Altman analysis. Data were analysed using GraphPad Prism 10.

## Results

### Analytical performance

Aptamer-AuNP-FcSH conjugates produced a strong and reversible Fc/Fc redox signal, confirming successful incorporation of the metallic redox reporter (**Fig S3B**). UV–visible spectra confirmed stepwise functionalisation, with the largest red-shift observed for co-functionalised nanoparticles (**Fig. S3A**). In spiked buffer samples with PCT, The ELLA-PCT generated a clear inverse relationship between PCT concentration and peak SWV current, consistent with the competitive binding format (**Fig. 3C**). The assay demonstrated an LOD of 46 pg/mL and a linear dynamic range of 0·5–100 ng/mL. Peak currents were stable at the established 30-minute time point, with coefficients of variation below 8%.

### Specificity, stability, and reproducibility

Testing with potential interferents at high concentrations showed minimal cross-reactivity. CRP, IL-6, and calcitonin produced apparent PCT values below 0·6 ng/mL, indicating low interference within clinically relevant thresholds (**Fig. 3D**). The assay also preserved its analytical performance after three months of storage at ambient temperature, with no detectable decline in redox peak current or in the slope of the calibration curve (**Fig. 3E**). ELLA-PCT showed good reproducibility in serum, with coefficients of variation of 4·99% for negative samples and 3·86– 10·74% across PCT-spiked serum samples ranging from 1 to 12 ng/mL (**Fig. 3F**), supporting the consistency of the assay across clinically relevant concentrations.

### Spiked Serum tests

In PCT-spiked serum samples, ELLA-PCT generated distinct electrochemical responses that reflected the competitive assay format, with lower PCT concentrations producing higher peak currents and higher PCT concentrations producing reduced signals (**Fig. 4A**). The calibration curve showed a clear inverse relationship between PCT concentration and peak current across the tested range, with a linear response against the logarithm of PCT concentration (**Fig. 4B**). This concentration-dependent behaviour was also visible on the lateral flow strips, where increasing PCT concentrations led to progressive fading of the test line before electrode insertion, confirming that the optical and electrochemical responses followed the same competitive binding mechanism (**Fig. 4C**).

**Figure 4.**
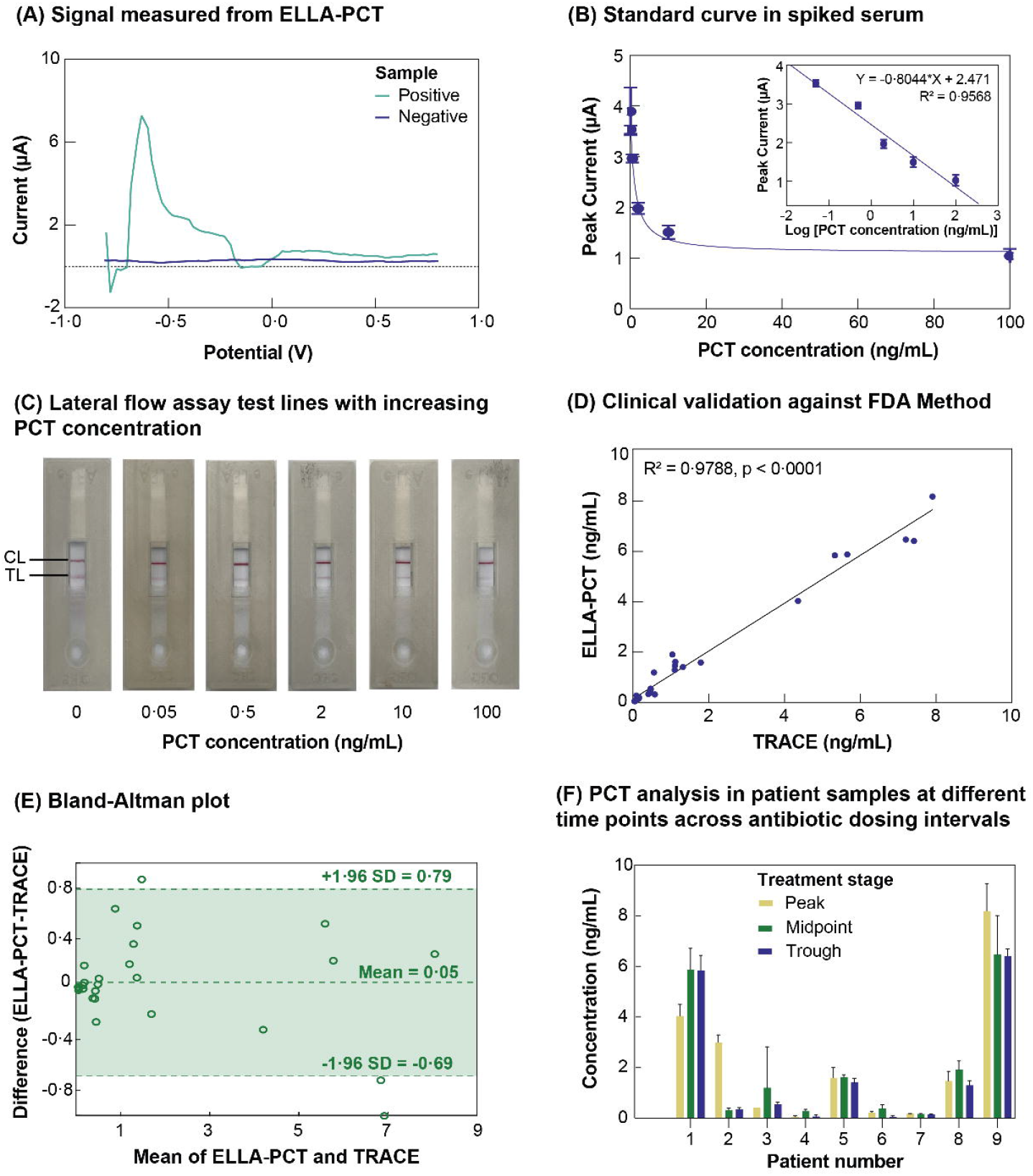
Analytical performance and clinical validation of ELLA-PCT in serum samples. **(A)** Representative square-wave voltammetry signals obtained from ELLA-PCT, showing the difference between high-current and low- current responses. Because ELLA-PCT uses a competitive assay format, lower PCT concentrations generate higher electrochemical currents, whereas higher PCT concentrations reduce the measured signal. **(B)** Calibration curve generated using PCT-spiked serum samples, showing the inverse relationship between PCT concentration and peak current. The inset shows the linear relationship between peak current and the logarithm of PCT concentration. (n = 3) **(C)** Representative lateral flow strips before electrode insertion, showing progressive fading of the test line with increasing PCT concentration. CL, control line; TL, test line. **(D)** Clinical validation of ELLA-PCT using 27 serum samples collected from nine patients undergoing antibiotic treatment, showing strong agreement with the TRACE reference method (R² = 0·9788, p < 0·0001). **(E)** Bland–Altman analysis comparing ELLA-PCT with TRACE, showing a mean bias of 0·05 ng/mL and 95% limits of agreement from –0·69 to 0·79 ng/mL, indicating good agreement between the two methods. **(F)** Analysis of clinical serum samples from nine patients receiving intravenous antibiotics for suspected bacterial infection. Three samples were collected per patient at predefined pharmacokinetic sampling timepoints relative to the antibiotic dosing interval: peak, midpoint, and trough. ELLA- PCT captured patient-to-patient variation in PCT concentrations across these real-world clinical serum samples (n = 3).

### Clinical performance

Across 27 serum samples, ELLA-PCT measurements showed strong correlation with the TRACE reference assay (R² = 0·979; p < 0·0001). Passing–Bablok regression (**Fig. 4D**) demonstrated a slope close to unity, and Bland–Altman analysis indicated a mean bias of 0.05 ng/mL with 95% limits of agreement between –0·69 and 0·89 ng/mL (**Fig. 4E**). In the clinical serum samples, ELLA-PCT captured variation in PCT concentrations across the patients and across predefined antibiotic dosing-interval sampling timepoints, including peak, midpoint, and trough samples (**Fig. 4F**). These data support proof-of-concept validation using real-world clinical samples, although the sampling design was not intended to evaluate longitudinal treatment response or infection resolution.

## Discussion

This study shows that a battery-free, smartphone-connected electrochemical lateral flow assay can provide quantitative PCT measurements in serum and clinical samples without conventional laboratory instrumentation. ELLA-PCT preserved the familiar workflow of an LFA while adding NFC-powered electrochemical readout, digital connectivity, and quantitative biomarker measurement. The assay generated PCT-dependent signals across clinically relevant concentrations, showed strong agreement with the TRACE reference method, and detected variation in PCT concentrations in serum samples from patients receiving antibiotics. These findings address an important gap between conventional LFAs, which are typically qualitative or semi-quantitative, and laboratory-based immunoassays, which provide quantitative results but require centralised infrastructure. In practice, decentralised access to quantitative PCT results could help healthcare professionals make faster antibiotic-treatment decisions, particularly around initiation, escalation, de-escalation, or discontinuation. Although demonstrated here for PCT, the ELLA architecture is modular: by changing the biorecognition element and capture chemistry, the same battery-free, smartphone-connected platform could be adapted to other clinically relevant biomarkers, including CRP, IL-6, serum amyloid A, or other inflammatory and host-response markers used in infection assessment.

The battery-free design is central to the practical value of the ELLA platform. Removing batteries reduces complexity of the device, avoids the need for charging or battery replacement, and improves suitability for storage and deployment in settings where access to power is limited. The ELLA platform also has an important environmental advantage: battery-free disposables reduce toxic waste and eliminate restrictions associated with battery-containing diagnostic devices. This is particularly relevant for high-volume point-of-care testing which are typically used in high volumes. By using the smartphone as the power source, ELLA-PCT retains digital connectivity while minimising the environmental footprint of the digital test.

The use of aptamers instead of antibodies provides an additional route towards scalable and robust point-of-care diagnostics. Aptamers are synthetic recognition elements that can offer improved stability, reproducible manufacture, and reduced dependence on cold-chain storage. These features are especially important for decentralised testing in outpatient clinics, emergency departments, care homes, and resource-limited settings including low- and middle-income countries. In this work, the aptamer-based format enabled an antibody-free assay for the detection of PCT while maintaining compatibility with the existing methods of manufacturing LFAs. The same approach could be adapted to other clinically relevant protein or peptide biomarkers with validated aptamers.

Despite these advantages, several limitations remain. The clinical evaluation was performed using a modest proof-of-concept cohort, and larger studies across diverse infection types, severity, and healthcare settings will be required to establish clinical performance with more confidence. The current validation was performed using serum samples, whereas whole-blood compatibility will be necessary for broader point-of-care implementation. Additional interference studies, assessment of operator-to-operator variability, and evaluation under real clinical workflow conditions will also be important. Furthermore, although ELLA-PCT could support antibiotic stewardship, interventional studies will be needed to determine whether implementation of the platform leads to measurable reductions in antibiotic use, treatment duration, or inappropriate prescription of antibiotics.

Future work should therefore focus on translating ELLA-PCT from analytical and early clinical validation toward workflow-integrated clinical testing. This includes adapting the assay for whole blood, expanding clinical validation, integrating the readout with electronic health records or antimicrobial stewardship platforms, and evaluating whether rapid access to quantitative PCT results impact clinical intervention. The ELLA architecture also provides a foundation for multiplexed point-of-care testing, where PCT could be combined with additional inflammatory, host-response, or pathogen-associated biomarkers to more accurately distinguish bacterial infection from viral disease, monitor treatment, or stratify patients at risk of deterioration.

In conclusion, ELLA-PCT demonstrates how a conventional LFA can be transformed into a quantitative, smartphone-connected digital diagnostic without incorporating batteries or sophisticated laboratory instrumentation. By combining quantitative electrochemical sensing, aptamer-based recognition, NFC-powered readout, and linked digital analytics, the platform provides a scalable template for clinical grade point-of-care diagnostics. Beyond PCT, the ELLA architecture could be extended to other biomarkers and application areas in which rapid, quantitative and decentralised testing is needed.

## Contributors

YC, JMRF, and FG conceived the study. YC and JMRF designed and performed the assay development and analytical experiments. AS designed and developed the mobile application for data acquisition, analysis, and user interfacing, and, together with TH and HSL, contributed to device engineering and PCB integration. LGM, ASC and HSL assisted with assay optimisation, manuscript revision and made the final figure SW, SF, RCW and TMR coordinated patient sample acquisition. RCW and TMR accessed and verified the underlying clinical data. FG supervised the project and provided critical revisions. All authors had access to the data, contributed to data interpretation, reviewed and approved the final manuscript, and accepted responsibility for submission.

## Data Availability

All data supporting the findings of this study are provided within the article and its supplementary materials.

## Data availability

All data supporting the findings of this study are provided within the article and its supplementary materials. The raw electrochemical voltammograms generated during the study are available in the accompanying Supplementary Data file.

## References

1. Rudd KE, Johnson SC, Agesa KM, et al. Global, regional, and national sepsis incidence and mortality, 1990–2017: analysis for the Global Burden of Disease Study. Lancet 2020; 395: 200–11.

2. Vijayan AL, Vanimaya, Ravindran S, et al. Procalcitonin: a promising diagnostic marker for sepsis and antibiotic therapy. J Intensive Care 2017; 5: 51.

3. Chambliss AB, Patel K, Colón-Franco JM, et al. AACC guidance document on the clinical use of procalcitonin. J Appl Lab Med 2023; 8: 598–634.

4. Samsudin I, Vasikaran SD. Clinical utility and measurement of procalcitonin. Clin Biochem Rev 2017; 38: 59–68.

5. Lee H. Procalcitonin as a biomarker of infectious diseases. Korean J Intern Med 2013; 28: 285–91.

6. Todd S, Euden J, Condie J, et al. Procalcitonin testing combined with NEWS2 evaluation compared with usual care based on NEWS2 for identification of sepsis and antibiotic initiation in the emergency department in England and Wales (PRONTO): a multicentre, randomised, controlled, open-label, phase 3 trial. Lancet Respir Med 2026; 14: 417–31.

7. Dark P, Hossain A, McAuley DF, et al. Biomarker-guided antibiotic duration for hospitalized patients with suspected sepsis: the ADAPT-Sepsis randomized clinical trial. JAMA 2025; 333: 682–93.

8. Prieto B, Llorente E, González-Pinto I, Alvarez FV. Plasma procalcitonin measured by time-resolved amplified cryptate emission (TRACE) in liver transplant patients: a prognosis marker of early infectious and non-infectious postoperative complications. Clin Chem Lab Med 2008; 46: 660–66.

9. Liao T, Yuan F, Yu H, Li Z. An ultrasensitive ELISA method for the detection of procalcitonin based on magnetic beads and enzyme-antibody labelled gold nanoparticles. Anal Methods 2016; 8: 1577–85.

10. Rieger M, Kochleus C, Teschner D, et al. A new ELISA for the quantification of equine procalcitonin in plasma as potential inflammation biomarker in horses. Anal Bioanal Chem 2014; 406: 5507–12.

11. Qi S, Li Q, Rao W, et al. Determining the concentration of procalcitonin using a magnetic particles-based chemiluminescence assay for the clinical diagnosis of sepsis. Anal Sci 2013; 29: 805–10.

12. Wang G, Wan Y, Lin G, et al. Development of a novel chemiluminescence immunoassay for the detection of procalcitonin. J Immunol Methods 2020; 484–485: 112829.

13. Jensch A, Mahla E, Toller W, et al. Procalcitonin measurement by Diazyme immunoturbidimetric and Elecsys BRAHMS PCT assay on a Roche COBAS modular analyser. Clin Chem Lab Med 2021; 59: e362–66.

14. Huang J, Zu Y, Zhang L, Cui W. Progress in procalcitonin detection based on immunoassay. Research 2024; 7: 0345.

15. Kinas BE, Akagac AE, Toprak AE, et al. Comparison of enzyme-linked fluorescent assay and electrochemiluminescence immune assay in procalcitonin measurement. Turk J Biochem 2022; 47: 19–22.

16. Dipalo M, Guido L, Micca G, et al. Multicenter comparison of automated procalcitonin immunoassays. Pract Lab Med 2015; 2: 22–28.

17. Ince B, Sezgintürk MK. Lateral flow assays for viruses diagnosis: up-to-date technology and future prospects. TrAC Trends Anal Chem 2022; 157: 116725.

18. Jiang N, Tansukawat ND, Gonzalez-Macia L, et al. Lateral and vertical flow assays for point-of-care diagnostics. Adv Healthc Mater 2019; 8: 1900244.

19. Dey MK, Iftesum M, Devireddy R, et al. New technologies and reagents in lateral flow assay (LFA) designs for enhancing accuracy and sensitivity. Anal Methods 2023; 15: 4351–76.

20. Gonzalez-Macia L, Li Y, Zhang K, et al. NFC-enabled potentiostat and nitrocellulose-based metal electrodes for electrochemical lateral flow assay. Biosens Bioelectron 2024; 251: 116124.

21. Olenik S, Lee HS, Güder F. The future of near-field communication-based wireless sensing. Nat Rev Mater 2021; 6: 286–88.

22. Flauzino JMR, Sanli A, Shirima RR, et al. Electrochemical lateral flow assay with ELISA- level performance for detecting plant diseases in East Africa. Proc Natl Acad Sci U S A 2026; 123: e2602947123.

23. Liu L, Li S, Liu J, et al. Aptamer-based biosensors for the diagnosis of sepsis. J Nanobiotechnology 2021; 19: 216.

24. Park DY, Shin WR, Kim SY, et al. In silico molecular docking validation of procalcitonin- binding aptamer and sepsis diagnosis. Mol Cell Toxicol 2023; 19: 843–55.

25. Nexha A, Niebuur BJ, Blum S, Kraus T. Agglomeration efficacies of simple salts on charged gold nanocrystals with mixed ligand shells: a high-throughput study. ACS Mater Au 2026; 6: 860–71.

26. Potts JC, Jain A, Amabilino DB, et al. Molecular surface quantification of multifunctionalized gold nanoparticles using UV-visible absorption spectroscopy deconvolution. Anal Chem 2023; 95: 12998–13002.

27. Göver T, Yazıcıgil Z. Electrochemical study of 6-(ferrocenyl)hexanethiol on gold electrode surface in non-aqueous media. Surf Interfaces 2018; 13: 163–67.

