## Supplementary Information for "Smartphone-connected, battery-free aptamer-based digital test for quantification of procalcitonin and monitoring of infection at the point of care"

**Supplementary Methods**

**Study design**

The study comprised four sequential phases: development and optimisation of the competitive aptamer-based electrochemical lateral flow assay; electrochemical characterisation of the signal probe and needle-electrode readout; analytical validation in buffer and serum; and proof-of-concept comparison with the hospital reference assay using longitudinal patient serum samples. Unless stated otherwise, experiments were performed at ambient laboratory temperature. The final assay was operated with 80 µL of sample and read 30 min after sample loading.

**Oligonucleotides and assay principle**

The PCT-binding DNA aptamer was a previously reported 51-nucleotide sequence: 5'-CCGCGGCAGTTCCGTAATGTTAATGCCTATACTTGAGCTGAGATAGTAAGT-3'. For nanoparticle conjugation, the aptamer was synthesised with a terminal thiol at the 5' end. The test line contained a biotinylated DNA strand complementary to the PCT aptamer and immobilised through a streptavidin-biotin bridge. The assay used a competitive format. In samples without PCT, the aptamer carried by the AuNP conjugate hybridised to the complementary strand at the test line, retaining ferrocene-labelled AuNPs and generating both a red colour and an electrochemical peak. When PCT was present, target binding reduced aptamer hybridisation at the test line; therefore, test-line colour and square-wave voltammetry (SWV) peak current decreased as PCT concentration increased. A separate anti-biotin AuNP population was captured by biotinylated BSA at the control line and served as a flow-control indicator.

**Validation of aptamer hybridisation and PCT recognition**

Aptamer-complement hybridisation was assessed by UV-visible spectroscopy. Equal amounts of the complementary strand or a non-complementary random strand were mixed separately with the aptamer-AuNP conjugate, and absorbance around 260 nm was compared with that of the single-stranded aptamer conjugate. Formation of the complementary duplex was indicated by the expected hypochromic decrease at 260 nm, whereas addition of the random strand increased absorbance because it remained predominantly single stranded. PCT recognition was also evaluated in a separate electrode-based experiment using an aptamer modified with a 5' thiol and a 3' ferrocene group. The aptamer was immobilised on a gold-plated needle electrode through the terminal thiol. A baseline SWV measurement was collected in PBS, after which the electrode was incubated for 10 min in PCT solutions of different concentrations, rinsed, and measured again in PBS. Concentration-dependent changes in the ferrocene peak current were used to confirm that the selected aptamer retained PCT-binding activity. This validation construct was used only to verify aptamer function and was not the signal architecture used in the final test.

**Salt-induced aggregation test for optimisation of aptamer loading**

A salt-induced aggregation screen was used to identify conditions that stabilised 40-nm AuNPs after aptamer immobilisation. The following buffers and pH values were evaluated: MES at pH 6.0 and 6.5, MOPS at pH 7.0 and 7.5, and borate buffer at pH 8.0, 8.5, and 9.0. Aptamer concentrations of 16, 32, 48, 64, and 80 µg/mL were tested. Controls comprised AuNPs challenged with 1 M NaCl, buffer plus 1 M NaCl without aptamer, and AuNPs without NaCl challenge. Following the salt challenge, nanoparticle aggregation was assessed by UV-visible absorbance, using the A550/A600 ratio as the stability metric. MOPS buffer at pH 7.0 and an aptamer concentration of 48 µg/mL were selected for preparation of the final conjugate.

**Preparation of aptamer-AuNP-FcSH and control conjugates**

One millilitre of 40-nm AuNP suspension at optical density 1 was mixed with 60 µL MOPS buffer (pH 7.0) and 30 µL of thiolated aptamer at 1.6 µg/µL, corresponding to 48 µg aptamer per millilitre of AuNP suspension. The suspension was gently mixed by inversion. Ferrocene hexanethiol (FcSH; 100 µL of a 10 mM solution in ethanol) was then added slowly while mixing by inversion. Addition of the aptamer before FcSH was selected because it produced the most stable conjugate and the strongest SWV response. The mixture was incubated for 24 h at room temperature to permit formation of Au-S bonds between the nanoparticle surface and both thiolated components. The conjugate was centrifuged at 5000 x g for 10 min, the supernatant was removed, and the pellet was resuspended in 250 µL of gold drying buffer.

A second AuNP colloid functionalised with goat anti-biotin antibody was used to generate the control line. Two hundred microlitres of the anti-biotin AuNP suspension were centrifuged at 5000 x g for 10 min. The supernatant was removed and the pellet was resuspended directly in the aptamer-AuNP-FcSH suspension, producing a combined conjugate mixture containing the PCT-responsive and flow-control particles. The final mixture was dispensed onto the conjugate pad and dried before strip assembly.

**Characterisation of functionalised AuNPs**

Bare AuNPs, AuNP-FcSH, AuNP-aptamer, and AuNP-aptamer-FcSH preparations were analysed by UV-visible spectroscopy. Stepwise surface functionalisation was evaluated from shifts and broadening of the AuNP surface-plasmon-resonance band relative to bare particles. FcSH-containing preparations were additionally assessed for increased absorbance in the approximately 320-400 nm region associated with the ferrocene moiety. Electrochemical activity of the conjugates was confirmed by drop-casting aliquots onto screen-printed gold electrodes and recording SWV. FcSH-functionalised particles were expected to generate a reversible Fc/Fc+ response. Ferrocene carboxylic acid (FcCOOH) was evaluated as a non-thiolated comparison compound; its substantially weaker electrochemical response supported selection of FcSH for the final conjugate.

**Preparation of test and control lines**

To anchor the nucleic-acid capture probe to nitrocellulose, equal volumes of streptavidin (1 mg/mL in PBS) and biotinylated complementary DNA (10 µM) were mixed and incubated for 2 h at 4 °C. This mixture was used to print the test line. The control-line reagent consisted of biotinylated BSA at 1 mg/mL in PBS containing 1% sucrose. Test- and control-line reagents were loaded into separate reservoirs of a BioDot dispensing system and applied to the nitrocellulose membrane as discrete lines, with the test line positioned upstream of the control line. Printed membranes were dried overnight at 37 °C.

**Fabrication and assembly of lateral-flow strips**

The strip comprised a glass-fibre sample pad, a glass-fibre conjugate pad containing the mixed AuNP conjugates, an FF170HP nitrocellulose detection membrane, and a cotton-linter absorbent pad mounted on an adhesive backing card (**Fig S1**). Adjacent components overlapped by approximately 2 mm to maintain continuous capillary flow. After drying, the components were aligned and laminated, and the assembled card was cut into strips 6 mm wide and approximately 60 mm long using an automated strip cutter. Each strip was inserted into a 3D-printed cassette designed to align the test line with the needle-electrode access points and the NFC potentiostat.

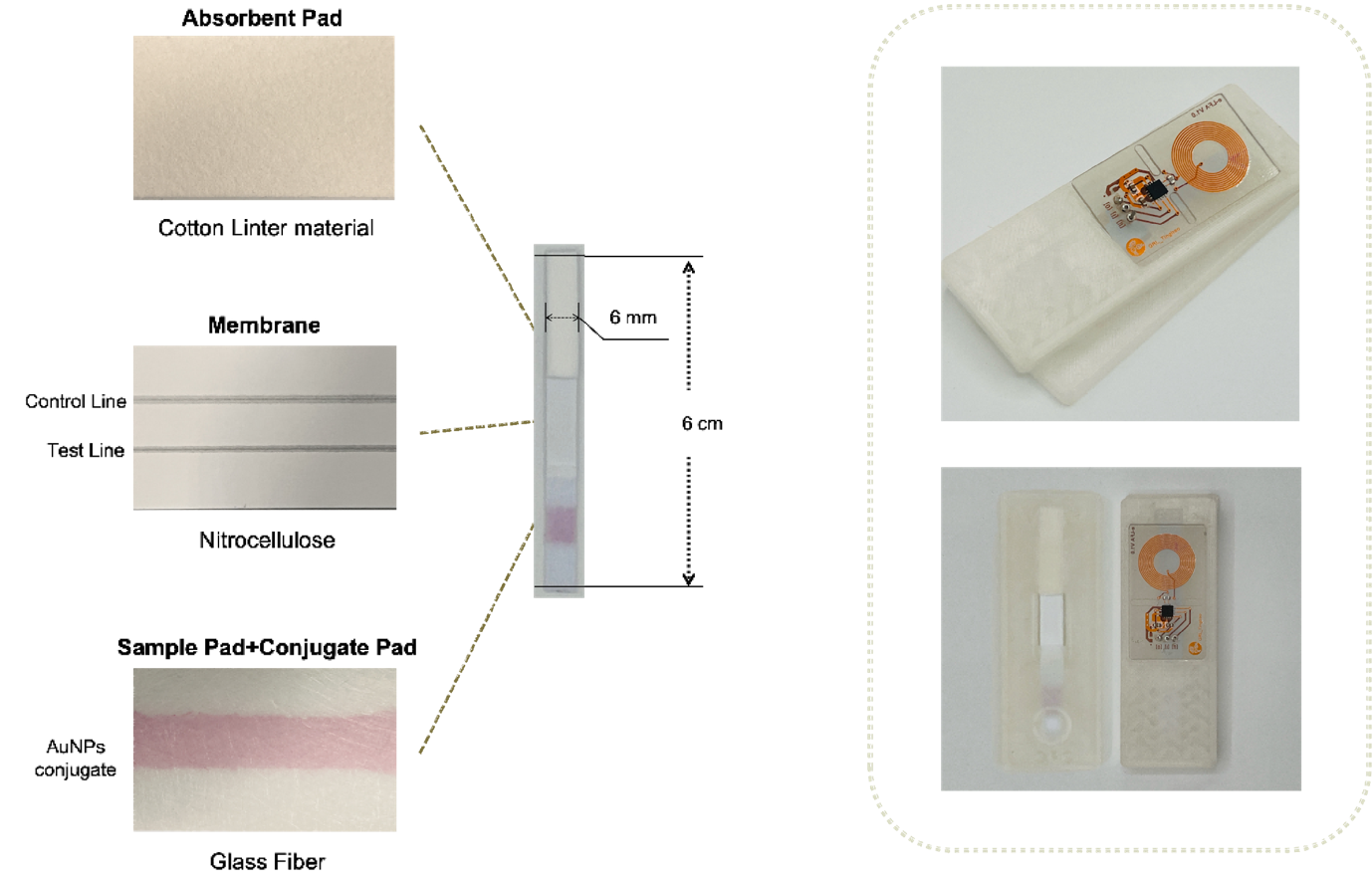

**Figure S1** – Scheme of the LFA construction and photos of the assembled eLFA.

**Optimisation of membrane, running buffer, and sample volume**

Two nitrocellulose membranes were compared: FF80HP, with a nominal capillary-flow time of 60-100 s, and FF170HP, with a nominal capillary-flow time of 140-200 s. Blank samples were used to compare maximum test-line formation. The slower-flowing FF170HP membrane produced the stronger line and was selected for subsequent experiments. Three candidate running buffers were compared: 150 mM PBS, 4 x SSC, and 100 mM Tris. Each buffer contained 2% (w/w) sucrose and 1% (v/v) Tween 20. The 4 x SSC formulation produced the strongest assay response and was selected as the final running buffer. Sample volumes of 20, 80, and 100 µL were then evaluated. Twenty microlitres did not sustain continuous flow, whereas 100 µL caused overflow from the strip and cassette. A volume of 80 µL provided complete migration without overflow and was used in all subsequent experiments. These optimisation experiments were initially assessed from the visual intensity and consistency of the test line.

**Needle electrodes and NFC potentiostat**

The electrochemical reader used a three-electrode acupuncture-needle array connected to a disposable printed circuit board (PCB). Gold-plated needles served as the working and counter electrodes, and a silver-plated needle served as the reference electrode. The PCB incorporated a SIC4341 NFC potentiostat (Silicon Craft Technology PLC, Thailand). When an NFC-enabled Android smartphone was positioned near the device, the phone provided wireless power, initiated the voltametric measurement, and received the electrochemical data without a battery or wired reader. A custom Android application displayed the voltammogram and the resulting PCT estimate. Electrochemical performance of the needle electrodes was evaluated using 5 mM ferrocyanide. SWV responses obtained with the gold-plated needles were compared with those from screen-printed gold electrodes. Needle-electrode stability was assessed over ten consecutive cyclic-voltammetry scans. The complete PCB-needle assembly was further tested by immersing the electrodes in 5 mM ferrocyanide and acquiring SWV through the smartphone application. This system-level test was performed using three independently assembled PCB-needle devices.

**Assay procedure and electrochemical measurement**

PCT standards or serum samples were prepared in the final running-buffer formulation (4 x SSC, 1% v/v Tween 20, and 2% w/w sucrose). An 80 µL aliquot was applied to the sample pad, and the strip was allowed to develop at ambient temperature. After 30 min, the three needles were inserted through the cassette so that they penetrated the wetted nitrocellulose at the test-line region. Electrodes were inserted only after chromatographic development to prevent the needles from disturbing capillary flow or damaging formation of the test line. SWV was recorded from -0.8 V to +0.8 V with a potential increment of 25 mV, an amplitude of 25 mV, a time step of 100 ms, and a frequency setting of 2.5 Hz. The Fc/Fc+ peak current generated by AuNP-aptamer-FcSH conjugates retained at the test line was extracted from each voltammogram. Because the assay was competitive, lower peak current indicated higher PCT concentration. The smartphone application received the SWV data by NFC and converted the test-line peak current to PCT concentration using the applicable calibration model.

**Optimisation of electrode-insertion time**

The effect of strip-development time was evaluated with a PCT-free running buffer, for which the maximum test-line current was expected. Separate strips were measured at 10, 15, 20, 25, 30, and 35 min after sample addition. Measurements at 10 min were unstable because the membrane and electrode-contact region were not fully equilibrated. Peak current increased between 15 and 30 min as the strip became uniformly wetted. At 35 min, the current decreased because the membrane had begun to dry. The 30-min time point produced the strongest stable response and was selected for all analytical and clinical measurements. Inserting the electrodes earlier and repeatedly recording the same developing strip was also evaluated, but this approach generated lower and less stable signals and was not used in the final protocol.

**Analytical calibration in buffer and serum**

For the buffer calibration, recombinant human PCT standards at 0, 0.05, 0.5, 2, 10, and 100 ng/mL were prepared in the running buffer. Each concentration was measured in triplicate using independently prepared strips, and the mean SWV peak current was plotted against PCT concentration. Additional calibration measurements were performed with PCT spiked into serum over the 0-100 ng/mL range; each serum calibration concentration was measured in triplicate. Calibration data were fitted using a four-parameter logistic model, with log-transformed concentration used for visualisation of the approximately linear portion of the response. Because increasing PCT produced a decreasing current, the analytical signal threshold for the limit of detection was defined as the mean current of the blank minus three standard deviations of the blank. This current threshold was converted to a PCT concentration using the fitted calibration equation, rather than being reported directly as a current value. The linear working range was defined from the portion of the calibration curve that maintained an approximately linear relationship between peak current and log10 PCT concentration.

**Interference, precision, and storage-stability studies**

Analytical specificity was assessed by testing potential interferers individually at concentrations exceeding their typical physiological ranges: CRP at 9.3 µg/mL, IL-6 at 2 ng/mL, and calcitonin at 100 ng/mL. Each interferent was processed using the same 80 µL, 30-min assay protocol. The measured current was converted to an apparent PCT concentration using the PCT calibration curve. Assay precision in serum was assessed using a negative serum sample and serum spiked with PCT at 1, 6, and 12 ng/mL. Peak-current coefficients of variation were calculated for each concentration. Storage stability was evaluated using strips stored at ambient temperature and tested after 1 day, 1 month, and 3 months. At each time point, strips were challenged with 0 and 100 ng/mL PCT, and the resulting peak currents were compared with those of freshly prepared strips.

**Clinical serum samples and reference testing**

The clinical proof-of-concept evaluation included 27 serum samples collected longitudinally from nine adults receiving antibiotic treatment for suspected bacterial infection. Three samples per participant were available across the treatment period. Each serum sample was analysed with the ELLA-PCT and by the hospital reference Time-Resolved Amplified Cryptate Emission (TRACE) assay. The ELLA-PCT operator obtained the electrochemical result using the same 80 µL loading volume and 30-min development time used for analytical validation. The study received approval from Imperial College London and the associated NHS research ethics committee, and all participants provided written informed consent.

**Statistical analysis**

Analytical calibration curves were fitted with a four-parameter logistic model. Precision was expressed as the coefficient of variation. Association between ELLA-PCT and TRACE measurements was assessed by regression analysis, including Passing-Bablok regression for method comparison. Agreement and systematic bias were evaluated using Bland-Altman analysis with the mean difference and 95% limits of agreement. Statistical analyses and graph generation were performed in GraphPad Prism version 10.

**Supplementary Results and Discussion**

**AuNPs aggregation test**

A salt-induced gold nanoparticle aggregation assay was used to identify conditions that maintained colloidal stability after aptamer immobilisation. Nanoparticle stability was assessed by UV-visible spectroscopy using the A550/A600 absorbance ratio. MOPS buffer at pH 7.0 and an aptamer concentration of 48 µg/mL provided the selected conditions for formation of the AuNP-aptamer conjugate (**Fig. S2**).

**Figure S2.** Absorbance ratio values (A₅₅₀/A₆₀₀) showing the stability of the gold nanoparticle conjugate in different concentrations of aptamer and buffers.
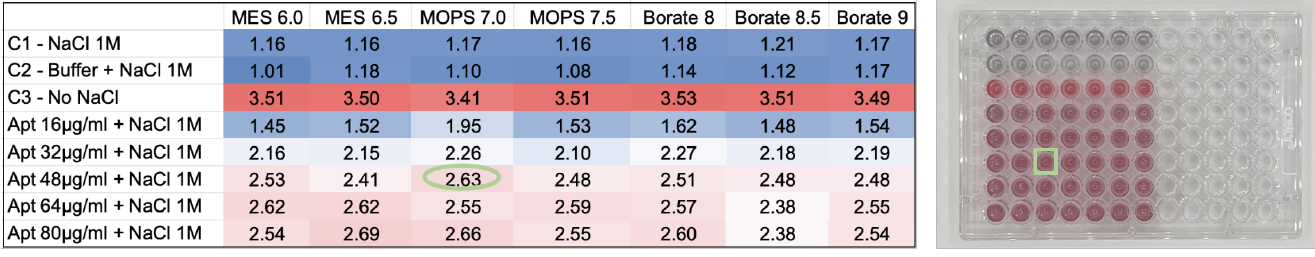

**AuNPs characterisation**

UV-visible spectroscopy showed progressive red-shifts and peak broadening after aptamer and FcSH modification of AuNPs, consistent with successful stepwise surface functionalisation (**Fig. S3A**). Importantly, SWV measurements of functionalised nanoparticles drop-cast onto screen-printed gold electrodes confirmed a clear Fc/Fc⁺ redox response for the FcSH-modified conjugates, whereas the non-thiolated FcCOOH control produced minimal electrochemical response (**Fig. S3B**). These results support the use of FcSH as a surface-anchored redox reporter capable of generating the electrochemical signal required for quantitative detection.

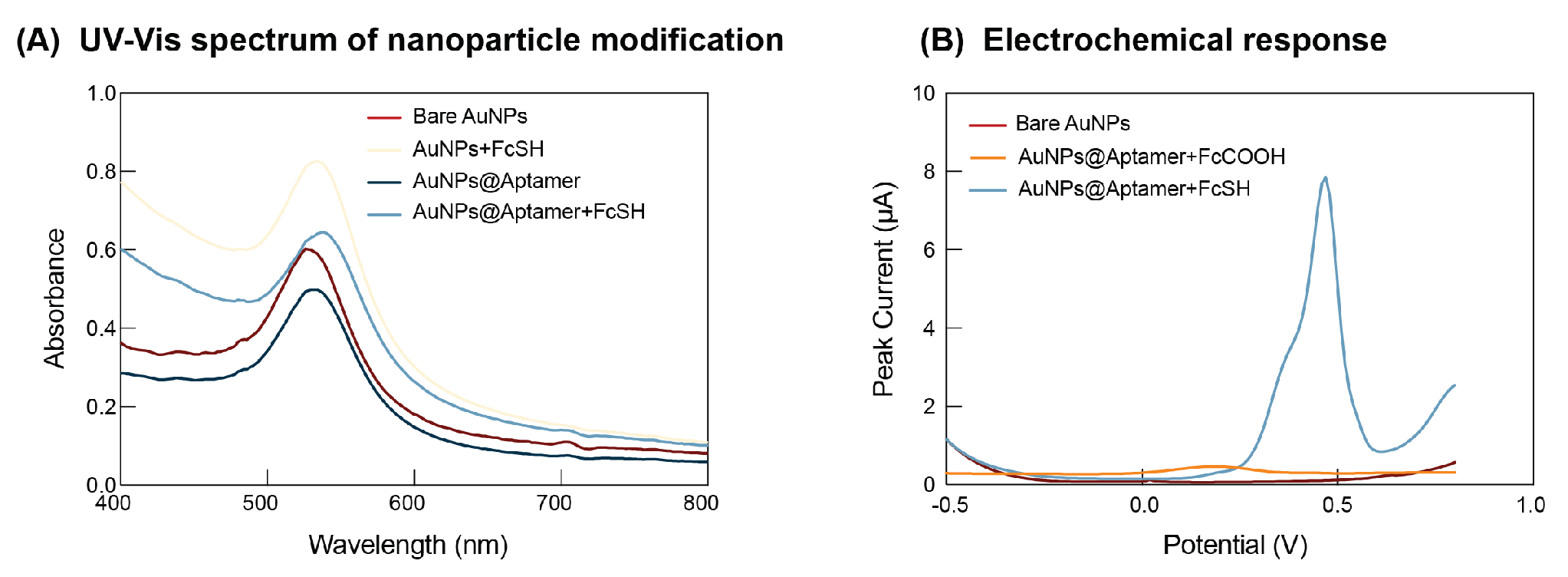

**Figure S3.** (A) UV-visible spectra showing progressive red-shifts and peak broadening following aptamer or FcSH modification, consistent with successful surface functionalisation. (B) SWV of functionalised nanoparticles drop-cast onto screen-printed gold electrodes, demonstrating Fc/Fc+ redox activity and confirming electrochemical responsiveness.

**Optimization of bioanalytical parameters of the ELLA-PCT**

Key lateral-flow parameters were optimised, including nitrocellulose membrane type, running-buffer composition, and sample volume. FF80HP and FF170HP membranes, with nominal capillary-flow times of 60-100 s and 140-200 s, respectively, were compared. The slower-flowing FF170HP membrane produced a stronger blank test line than FF80HP, consistent with increased interaction time between the conjugate and the immobilised complementary strand. Three running buffers were evaluated: 150 mM PBS, 4 x SSC, and 100 mM Tris, each supplemented with 2% (w/w) sucrose and 1% (v/v) Tween 20. The 4 x SSC formulation produced the strongest response and was selected for the final assay. Sample volumes of 20, 80, and 100 µL were also compared. Twenty microlitres did not sustain complete flow, whereas 100 µL caused overflow. A volume of 80 µL supported stable migration and a strong signal without overflow. Initial selection of these parameters was based on visual test-line intensity and consistency.

**Optimisation of needle-electrode insertion time**

Needle-electrode insertion time was evaluated using PCT-free running buffer, which generated the maximum test-line signal in the competitive assay. Separate strips were measured at 10, 15, 20, 25, 30, and 35 min after sample addition. Measurements at 10 min were unstable because the strip and electrode-contact region were not fully saturated. Signal increased between 15 and 30 min as the membrane equilibrated, whereas insertion at 35 min produced a lower current as the strip began to dry. The 30-min time point provided the strongest stable response and was selected for subsequent measurements (**Fig. S4**).

**Figure S4.** Optimisation of needle-electrode insertion time: (A) SWV peak currents measured at different time points; (B) relationship between peak current and insertion time from 15 to 30 min; and (C) visual appearance of strips at representative time points.
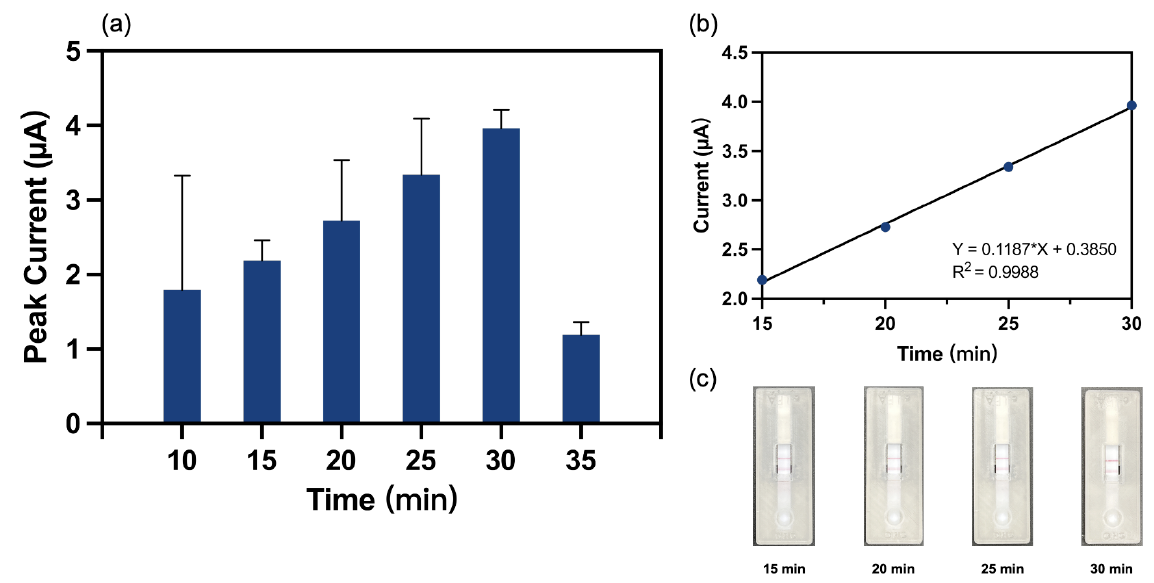

**Electrode characterisation**

The electrochemical readout architecture was further validated by comparing the gold-plated acupuncture-needle electrodes with conventional screen-printed gold electrodes in ferrocyanide solution. The needle electrodes generated comparable redox responses, confirming their suitability for on-strip electrochemical interrogation (**Fig. S5A**). In addition, the gold-plated needles maintained stable cyclic-voltammetry responses over the first five consecutive scans, indicating that the electrode surface remained electrochemically stable during repeated measurements (**Fig. S5B**). After five voltametric cycles, the current response at higher potentials began to increase, probably due to partial removal of the thin gold layer from the needles. This effect is unlikely to affect ELLA-PCT performance because the device is designed for single-use operation, and only one voltametric scan is required for sample detection. Together, these results support the use of disposable gold-plated needle electrodes as a practical alternative to conventional electrodes for integration into the battery-free ELLA-PCT cassette.

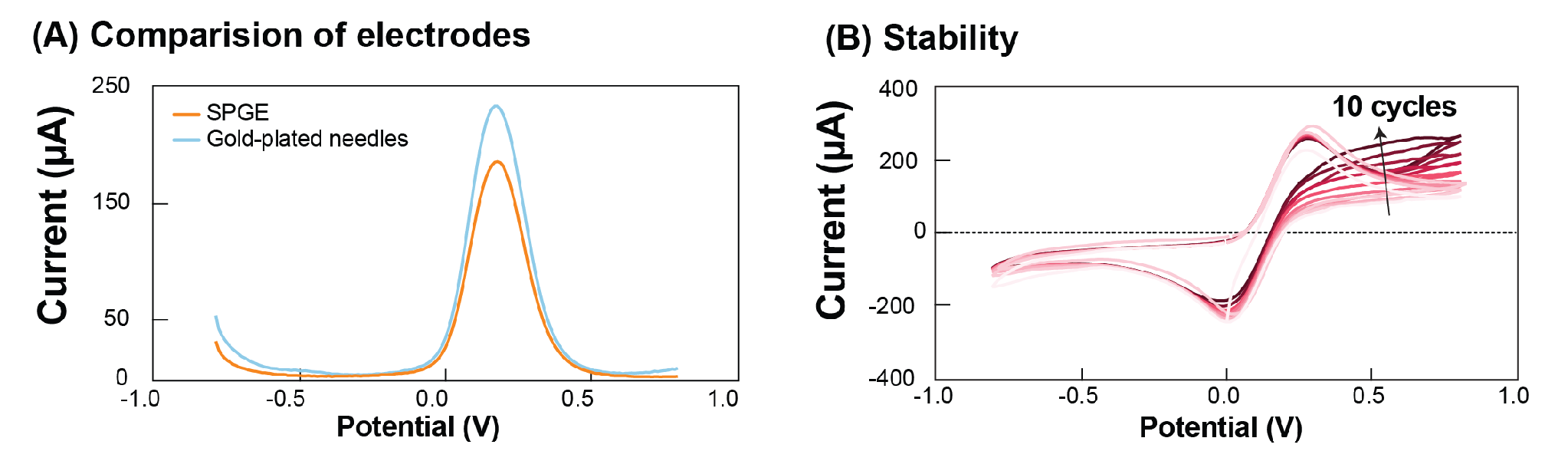

**Figure S5.** (A) SWV comparison between screen-printed gold electrodes and gold-plated needle electrodes in ferrocyanide solution. (B) Stability of the gold-plated needle electrodes during ten consecutive cyclic-voltammetry cycles in a redox-probe solution.

**Comparison with other procalcitonin biosensors in the literature**

**Table S1 –** Comparison of different biosensors for procalcitonin detection.

| **Biosensor Type** | **Detection Method** | **LOD** | **Linear Range** | **Ref.** |
| --- | --- | --- | --- | --- |
| ECL-RET immunosensor (g-C3N4-AuD@PPy/ZnONFs@PDA-sCuO) | Electrochemiluminescence Resonance Energy Transfer | 17.2 fg/mL | 0.00005-50 ng/mL | ^1^ |
| Eu-MOF + CoS2 TSNBs-based ECL immunosensor | Near-infrared Electrochemiluminescence | 3.65 fg/mL | 10 fg/mL-100 ng/mL | ^2^ |
| CdS/BiVO4/GaON-based PEC immunosensor | Photoelectrochemical Detection | 0.03 pg/mL | 0.1 pg/mL-50 ng/mL | ^3^ |
| Plasmonic imaging digital immunoassay | Time-Resolved Plasmonic Imaging | 2.8 pg/mL | 4.2-12,500 pg/mL | ^4^ |
| LFA with AuNP and electrochemical readout | Colorimetric + Cyclic Voltammetry | 1 ng/mL (quantitative) | Not specified | ^5^ |
| PEDOT:PSS-AuNP on CFP | Electrochemical (Chronoamperometry) | 1 pg/mL | 1-200,000 pg/mL | ^6^ |
| Cu-BHT thin film sensor | Electrochemical Impedance Spectroscopy | 14.6 pg/mL | 0.1-100 ng/mL | ^7^ |
| NH2-VMSF with in-situ AuNPs | Electrochemiluminescence (Luminol-based) | 7 pg/mL | 10 pg/mL-100 ng/mL | ^8^ |
| WENLISA (Waveguide-Enhanced Nanogold-Linked Immunosorbent Assay) | Optical Plasmonic Waveguide Absorption | 48.7 fg/mL | 0.1 pg/mL-1 ng/mL | ^9^ |
| ELLA-PCT | Electrochemical (Square Wave Voltammetry) | 0.046 ng/mL | 0.5-100 ng/m | This work |

**References**

1.	Shi, T. *et al.* Dual-quenching electrochemiluminescence system based on resonance energy transfer from gold dendrite@polypyrrole core–shell nanoparticles enhanced g-C3N4 to ZnONFs@PDA-sCuO for procalcitonin immunosensing. *Sens Actuators B Chem* **371**, 132591 (2022).

2.	Zhao, L. *et al.* Ultrasensitive near-infrared electrochemiluminescence biosensor derived from Eu-MOF with antenna effect and high efficiency catalysis of specific CoS2 hollow triple shelled nanoboxes for procalcitonin. *Biosens Bioelectron* **191**, 113409 (2021).

3.	Li, S. *et al.* A sensitive biosensor of CdS sensitized BiVO4/GaON composite for the photoelectrochemical immunoassay of procalcitonin. *Sens Actuators B Chem* **329**, 129244 (2021).

4.	Jing, W. *et al.* Time-Resolved Digital Immunoassay for Rapid and Sensitive Quantitation of Procalcitonin with Plasmonic Imaging. *ACS Nano* **13**, 8609–8617 (2019).

5.	Gupta, Y., Kalpana & Ghrera, A. S. Electrochemical studies of lateral flow assay test results for procalcitonin detection. *Journal of Electrochemical Science and Engineering* **12**, 265–274 (2022).

6.	Gupta, Y. & Ghrera, A. S. Development of conducting paper-based electrochemical biosensor for procalcitonin detection. *ADMET DMPK* **11**, 263–275 (2023).

7.	Guo, Z. *et al.* An ultra-sensitive electrochemical biosensor for the detection of procalcitonin in sepsis patients’ serum, using a Cu-BHT-based thin film. *Talanta* **268**, 125325 (2024).

8.	Chang, Q., Gu, X., He, L. & Xi, F. A highly sensitive immunosensor based on nanochannel-confined nano-gold enhanced electrochemiluminescence for procalcitonin detection. *Front Chem* **11**, 1274424 (2023).

9.	Barshilia, D. *et al.* Ultrasensitive and Rapid Detection of Procalcitonin via Waveguide-Enhanced Nanogold-Linked Immunosorbent Assay for Early Sepsis Diagnosis. *Nano Lett* **24**, 2596–2602 (2024).
